# Schools are not islands: Balancing COVID-19 risk and educational benefits using structural and temporal countermeasures

**DOI:** 10.1101/2020.09.08.20190942

**Authors:** Jamie A. Cohen, Dina Mistry, Cliff C. Kerr, Daniel J. Klein

## Abstract

**Background:** School closures around the world contributed to reducing the transmission of COVID-19. In the face of significant uncertainty around the epidemic impact of in-person schooling, policymakers, parents, and teachers are weighing the risks and benefits of returning to in-person education. In this context, we examined the impact of different school reopening scenarios on transmission within and outside of schools and on the share of school days that would need to be spent learning at a distance.

**Methods:** We used an agent-based mathematical model of COVID-19 transmission and interventions to quantify the impact of school reopening on disease transmission and the extent to which school-based interventions could mitigate epidemic spread within and outside schools. We compared seven school reopening strategies that vary the degree of countermeasures within schools to mitigate COVID-19 transmission, including the use of face masks, physical distancing, classroom cohorting, screening, testing, and contact tracing, as well as schedule changes to reduce the number of students in school. We considered three scenarios for the size of the epidemic in the two weeks prior to school reopening: 20, 50, or 110 detected cases per 100,000 individuals and assumed the epidemic was slowly declining with full school closures (*R*_e_ = 0.9). For each scenario, we calculated the percentage of schools that would have at least one person arriving at school with an active COVID-19 infection on the first day of school; the percentage of in-person school days that would be lost due to scheduled distance learning, symptomatic screening or quarantine; the cumulative infection rate for students, staff and teachers over the first three months of school; and the effective reproduction number averaged over the first three months of school within the community.

**Findings:** In-person schooling poses significant risks to students, teachers, and staff. On the first day of school, 5–42% of schools would have at least one person arrive at school with active COVID-19, depending on the incidence of COVID in the community and the school type. However, reducing class sizes via A/B school scheduling, combined with an incremental approach that returns elementary schools in person and keeps all other students remote, can mitigate COVID transmission. In the absence of any countermeasures in schools, we expect 6 – 25% of teaching and non-teaching staff and 4 – 20% of students to be infected with COVID in the first three months of school, depending upon the case detection rate. Schools can lower this risk to as low as 0.2% for staff and 0.1% for students by returning elementary schools with a hybrid schedule while all other grades continue learning remotely. However, this approach would require 60–85% of all school days to be spent at home. Despite the significant risks to the school population, reopening schools would not significantly increase community-wide transmission, provided sufficient countermeasures are implemented in schools.

**Interpretation:** Without extensive countermeasures, school reopening may lead to an increase in infections and a significant number of re-closures as cases are identified among staff and students. Returning elementary schools only with A/B scheduling is the lowest risk school reopening strategy that includes some in-person learning.

**Research in context:** 

**Evidence before this study:** Scientific evidence on COVID-19 transmission has been evolving rapidly. We searched PubMed on 6 September 2020 for studies using the phrase (“COVID-19” OR “SARS-CoV-2”) AND (“model” OR “modeling” OR “modelling”) AND (“schools”) AND (“interventions”). This returned 17 studies, of which 6 were retained after screening. A wide variety of impacts from school closures were reported: from 2–4% of deaths at the lower end to reducing peak numbers of infections by 40–60% at the upper end. Drivers of this variability include (a) different epidemic contexts when school closure scenarios are enacted, (b) different timeframes and endpoints, and (c) different model structures and parameterizations. Thus, considerable variation in predicted impacts of school closures has been reported.

**Added value of this study:** To our knowledge, this is the first modeling study that explores the trade-offs between increased risk of COVID-19 transmission and school days lost, taking into account detailed data on school demographics and contact patterns, a set of classroom countermeasures based on proposed policies, and applies them to range of community transmission levels. If rates of community transmission are high, school reopening will accelerate the epidemic, but will not change its overall course. However, even if rates of community transmission are low, complete school reopening risks returning to exponential epidemic growth. Staged school reopening coupled with aggressive countermeasures is the safest strategy, but even so, reactive school closures will likely be necessary to prevent epidemic spread.

**Implications of all the available evidence:** The impact of school reopening on the COVID-19 epidemic depends on the transmission context and specific countermeasures used, and no reopening strategies are zero risk. However, by layering multiple types of countermeasures and responding quickly to increases in new infections, the risks of school reopening can be minimized.

## 1. Introduction

Primary and secondary schools around the world closed due to COVID-19 transmission in March and April 2020. A recent observational study [1] estimated that school closures were associated with a 62% decline in weekly COVID-19 incidence and a 58% decline in weekly COVID-19 mortality; some modeling studies have also found large impacts of school closures [18, 23]. However, several other studies [29, 9, 22] have estimated that closing schools has only a relatively small impact (e.g., 2–4% reduction in deaths).

Attention is now focused on the risks and benefits of reopening schools. In-person learning has educational, social, and physical benefits; schools provide a safe space for educational instruction, support the development of social and emotional skills, and address nutritional needs, especially for families of lower socioeconomic status. For these reasons and more, the American Academy of Pediatrics issued a statement [25] on July 10 advocating for bringing students back to the classroom for in-person learning.

Yet much remains uncertain about the role schools, and school-age students in particular, play in COVID-19 transmission, as well as how effective school-based interventions will be in preventing transmission. While children may be both less susceptible and less likely to develop severe infection, nearly a quarter of teachers are at a greater risk of serious illness if infected with COVID-19 [6]. In an NPR/Ipsos poll of 505 teachers across the US conducted in July, 77% of teachers indicated that they are worried about risking their own health in returning to in-person education and 66% would prefer primarily remote, distance learning [15].

Much can be learned from the experience of school reopening across Europe and Asia; some countries successfully reopened without a significant increase in COVID-19 cases [13], while others were forced to quickly re-close after outbreaks were observed in multiple schools [7, 28]. In general, countries that reopened schools tended to do so when the case detection rate over the most recent 14 day period was below 25 per 100,000 – nearly 10 times lower than the case detection rate in the United States as of August 11 [20], with significant heterogeneity both across and within states. Additionally, many of these settings returned younger students in-person first with some degree of staggering the start, stop, and break times within schools.

In this analysis, we used an agent-based model of COVID-19 transmission to evaluate the health and educational outcomes associated with various school reopening strategies. Specifically we compared behavioral interventions such as mask usage, physical distancing and hand hygiene; case detection interventions, including symptomatic screening, follow-up diagnostic testing and contact tracing; and structural interventions, including classroom cohorting, schedule changes, and combinations of remote and in-person learning.

## 2. Methods

### 2.1. Model overview

We used Covasim, an agent-based model of COVID-19 transmission and interventions [16, 17], to estimate the impact of school reopening on disease transmission and the extent to which school-based interventions could mitigate epidemic spread within and outside schools. Covasim includes demographic information on age structure and population size; realistic transmission networks in different social layers, including households, schools, workplaces, long-term care facilities and communities; age-specific disease outcomes; and within- and between-host variations in infectivity to capture sub- and super-spreading and front-loaded infectivity. We modeled schools to match age mixing patterns between students within elementary schools, middle schools, and high schools [10] (see Appendix A for more details on the school network structures). Key inputs and assumptions of our modeling approach have been documented in our methodology papers [16, 17]. The Covasim model is fully open source and is available for download via GitHub and the Python Package Index; more information is available at https://covasim.org.

### 2.2. School reopening scenarios

We compared seven alternative school reopening strategies that vary the degree of school-based transmission mitigation. Countermeasures were non-pharmaceutical interventions (NPIs), which implicitly include face masks, six foot separation, and hand washing, which together are assumed to reduce transmission by 25% [5, 19, 4]; class cohorting, in which students and teachers have minimal contact outside their own classroom (see Figure 1); and symptomatic screening, with 50% follow-up diagnostic testing and 50% follow-up contact tracing. A/B scheduling splits classrooms into group A students (who attend two days per week, e.g. Mon-Tue) and group B (who attend two different days per week, e.g. Thu-Fri).

1. All in-person with no countermeasures
2. All in-person with countermeasures
3. All in-person with countermeasures and A/B scheduling
4. Elementary and middle in-person with countermeasures, high school remote
5. Elementary in-person with countermeasures, middle and high school remote
6. Elementary in-person with countermeasures and A/B scheduling, middle and high school remote
7. All remote

**Figure 1:**
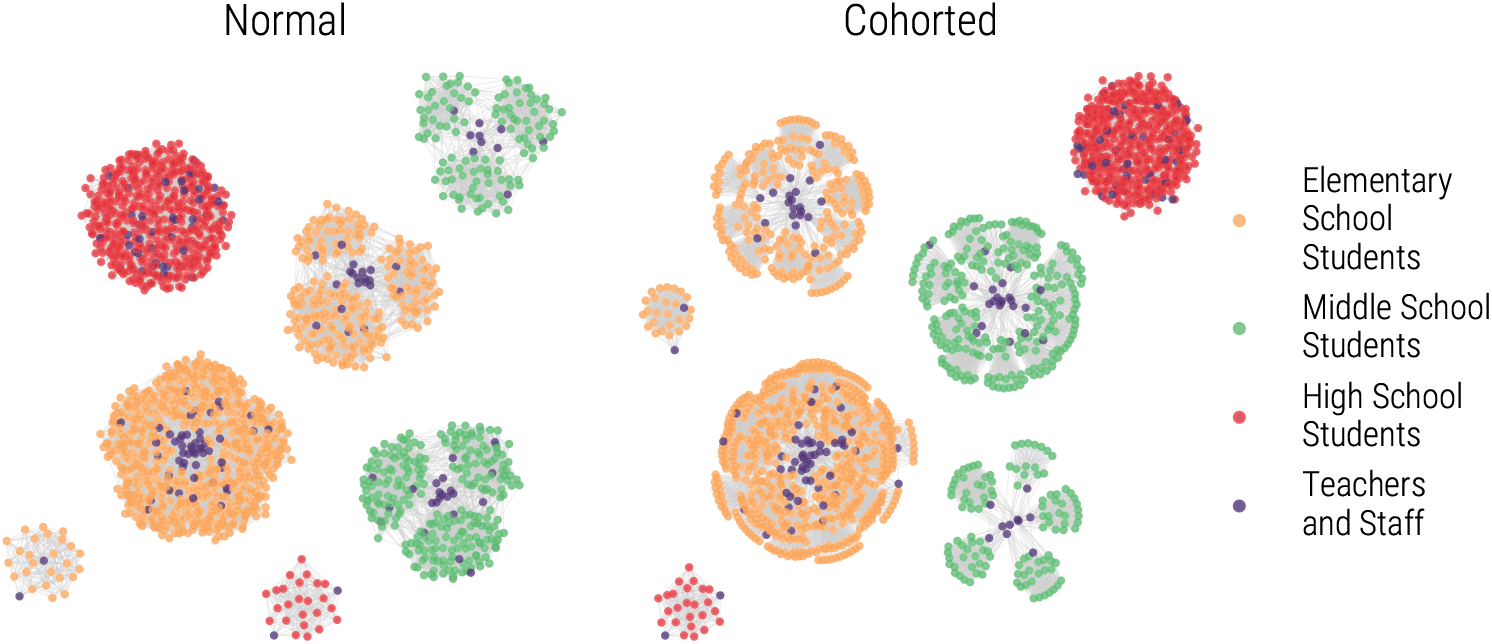
A schematic diagram of the in-person school contact networks of students, teachers and additional school staff under different cohorting strategies. Normal, pre-COVID-19 patterns allow for mixing of students and teachers across grades and classrooms. A cohorted strategy places elementary and middle school students into classrooms with their own teachers, preventing contact between students in different classrooms in these schools. High schools remain mixed due to the highly individualized schedules of students in many U.S. based high schools, including those in King County, Washington. Teachers and additional school staff have contacts with each other to reflect their use of common staff paces such as a teachers’ lounge, front office, and other rooms for preparation.

We applied our interventions to elementary, middle, and high schools and assumed that preschools and universities will remain closed. We assumed that high schools would not be able to implement classroom cohorting, as it would be too challenging to coordinate the highly variable schedules of students at this level (see Figure 1 for a schematic of the school network differences). We simulated the first three months of the school term (Sept. 1^st^-Dec. 1^st^).

We used data from King County, Washington to define the population and contact network structure, including the average class size and the distribution of students enrolled per school [26]. The effective reproduction number in the absence of school reopening and the case detection rate in the two weeks prior to school reopening served as signals of both the direction of cases as well as the size of the epidemic. Results presented in this work assume that epidemic is declining slowly when schools are fully closed (i.e., *R*_e_ = 0.9). In conjunction with our assumption around epidemic control with full school closures, we considered three scenarios for the size of the epidemic in the two weeks prior to school reopening: 20, 50, or 110 diagnosed cases per 100,000 individuals. These numbers fit within the low, medium and high case detection bands defined by states across the United States for school re-opening guidance.

## 3. Results

In-person schooling, even with sufficient countermeasures, poses significant risks to students, teachers, and staff. On the first day of school, 5 – 42% of schools would have at least one person arrive at school with active COVID-19 (including all students, teachers, and staff), depending on the COVID case detection rate in the community and the school type (Figure 2). These infections may show few symptoms and go undetected, especially if they are in younger children. Symptomatic individuals may be screened and sent home immediately upon arrival. Active COVID-19 infections also may not lead to onward transmission within schools, depending on per-contact infectivity. This highlights the importance of procedures within schools to minimize risk of transmission, detect and isolate cases, and contact and quarantine any known contacts.

**Figure 2:**
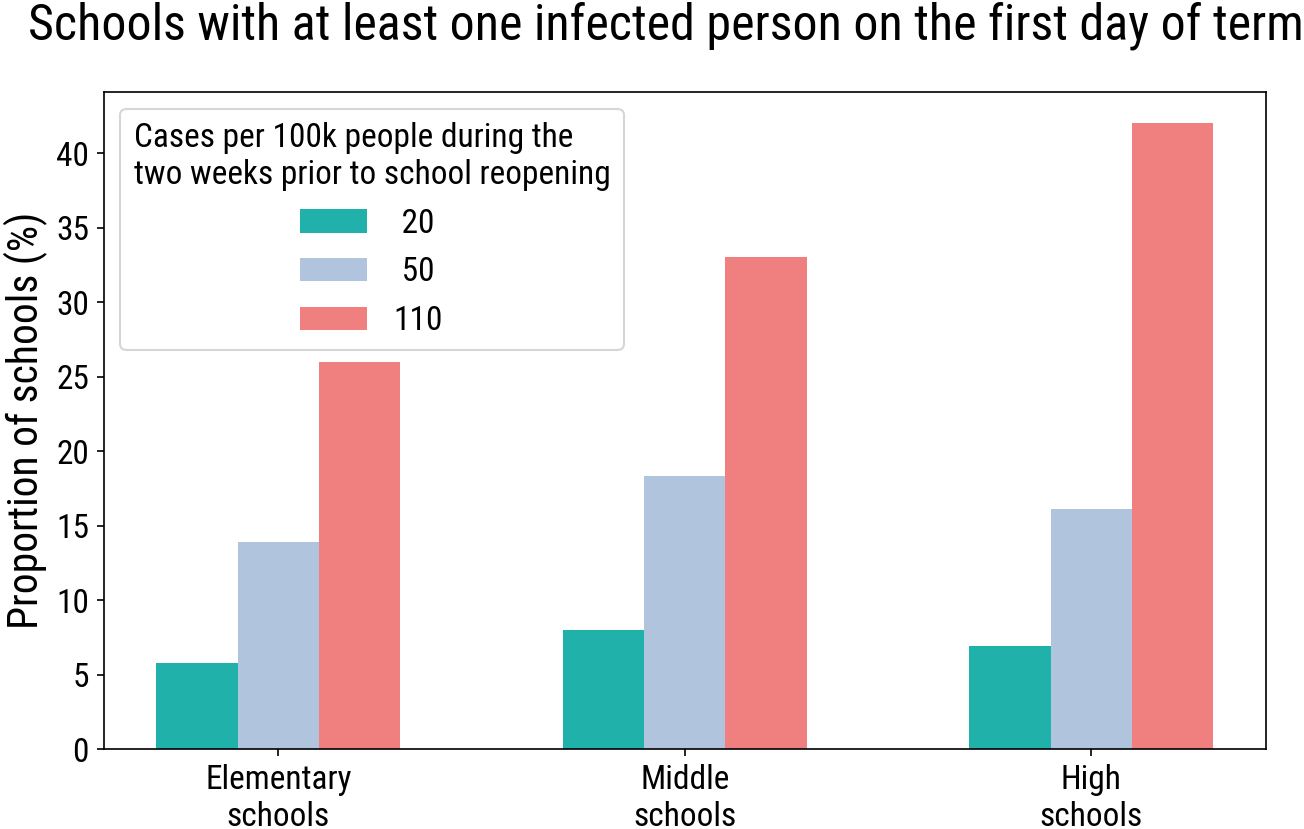
Percent of schools with at least one infectious individual on the first day of school, averaged across the top 20 parameter sets.

Closely examining infections present in school, our model shows that in the absence of any countermeasures and at a case detection rate of 110 per 100,000 in the two weeks prior to school reopening, nearly a quarter of teachers and staff and over 17% of students would visit school while infectious with COVID-19 in the first three months of school (Figure 3). This risk can be reduced 2.5-fold by reopening schools when the case detection rate is 20 per 100,000; when there is more COVID in the population, there will inevitably be more COVID in schools.

**Figure 3:**
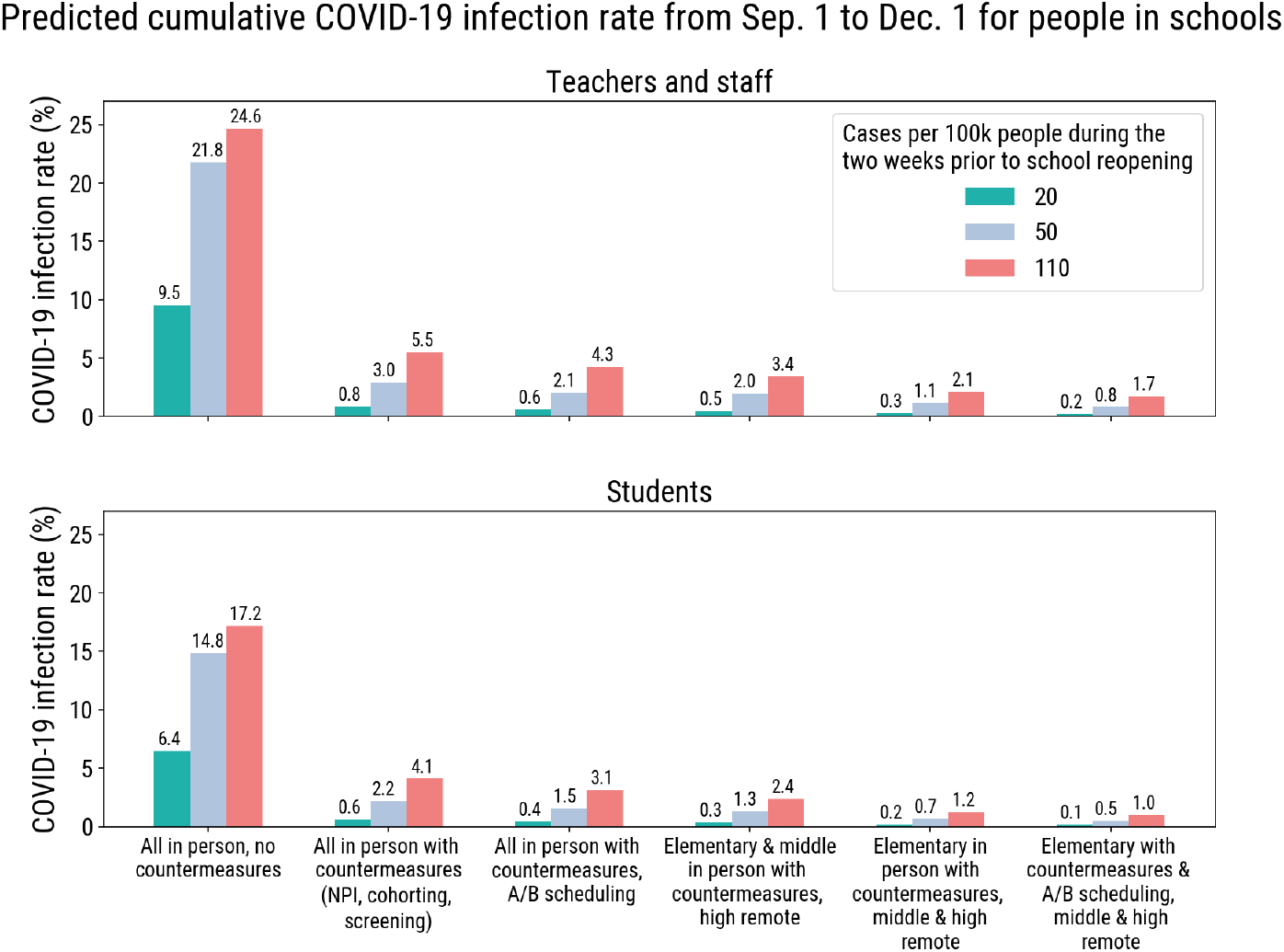
Cumulative COVID-19 infection rate for students, teachers and staff physically present in schools during the period of school reopening (Sept 1^st^-Dec 1^st^), averaged across the top 20 parameter sets.

We find that implementing countermeasures that limit transmission and detect, trace, and quarantine cases within schools would lead to an even larger reduction in the cumulative COVID-19 infection rate at each of the three case detection rates we considered. The inclusion of face mask usage, physical distancing, classroom cohorting, symptomatic screening, testing and tracing in schools would lower the risk to students, teachers, and staff over 4-fold at a case detection rate of 110 per 100,000. This risk can be lowered further with the addition of scheduling efforts that reduces the number of students present at school. An A/B school scheduling approach that returns elementary schools in-person and keeps all other students remote would minimize the presence of COVID within (and outside of) schools. The predicted cumulative COVID-19 infection rate for people in schools could be as low as between 0.2 and 1.7% for teachers and staff and to between 0.1 and 1.0% for students, depending on the case detection rate. This represents at least a 14-fold reduction in the risk of COVID-19 for teachers and staff in schools relative to a strategy that returns all individuals in-person with no countermeasures. Within the King County population of approximately million, we estimate that this approach would require an additional 900– 6,200 tests for students, teachers and staff over the first three months of school, depending on the case detection rate in the two weeks prior to school.

We find that an A/B scheduling approach, in which classrooms are split into two groups that attend school two days a week on different days, reduces COVID-19 transmission in schools to 0.6 – 4.3% for teachers and staff and 0.4– 3.1% for students, depending upon the COVID case detection rate. This represents about double the risk compared to an elementary-only approach, but gives all K-12 students some time for in-person learning, whereas the elementary-only approach restricts in-person learning to elementary school students, at least initially.

These strategies come at a significant educational cost, requiring up to 83% of school days to be spent at home, due to either planned distance learning or related to detected COVID-19 infection (Figure 4). Provided sufficient countermeasures are implemented within schools, the COVID-19 infection rate in the population prior to school reopening has more influence on the infection rate within schools than the specific schooling strategy. We find an over seven times reduction in the infection rate for people in schools if schools are reopened when the case detection rate in the community is at 20 per 100,000 compared to 110 per 100,000. At any given case detection rate, additional countermeasures will have a marginal impact on the rate of infection within schools at a large cost of missed in-person school days.

**Figure 4:**
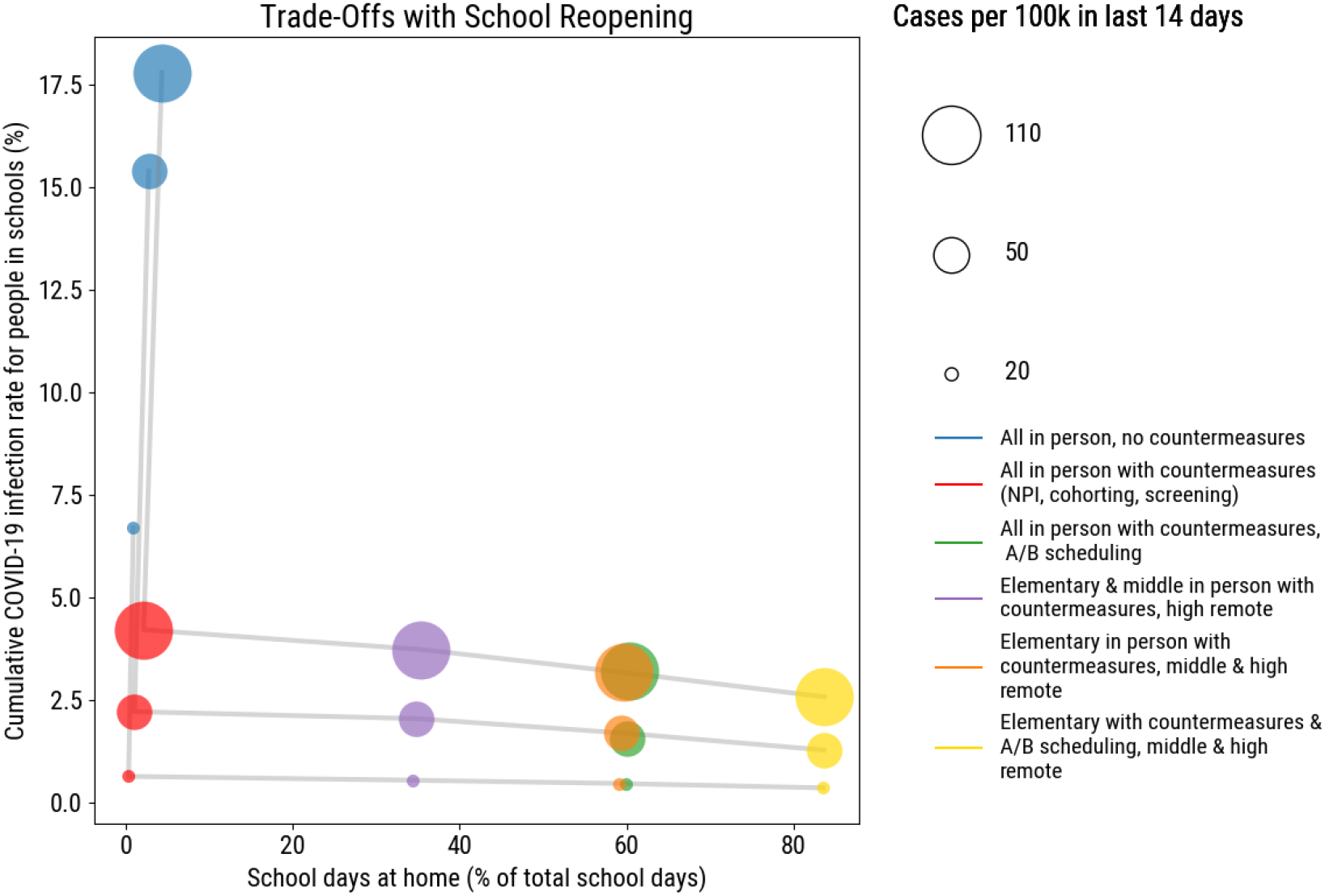
Trade-offs between within-school infection rate and missed in-person school days (due to either scheduled distance learning, quarantine or infection.

We find that reopening schools will not significantly increase community-wide transmission, provided sufficient school-based interventions are implemented (Figure 5). If community transmission is decreasing in the absence of in-person schooling, a return to in-person learning with appropriate countermeasures is unlikely to add significantly to community transmission.

**Figure 5:**
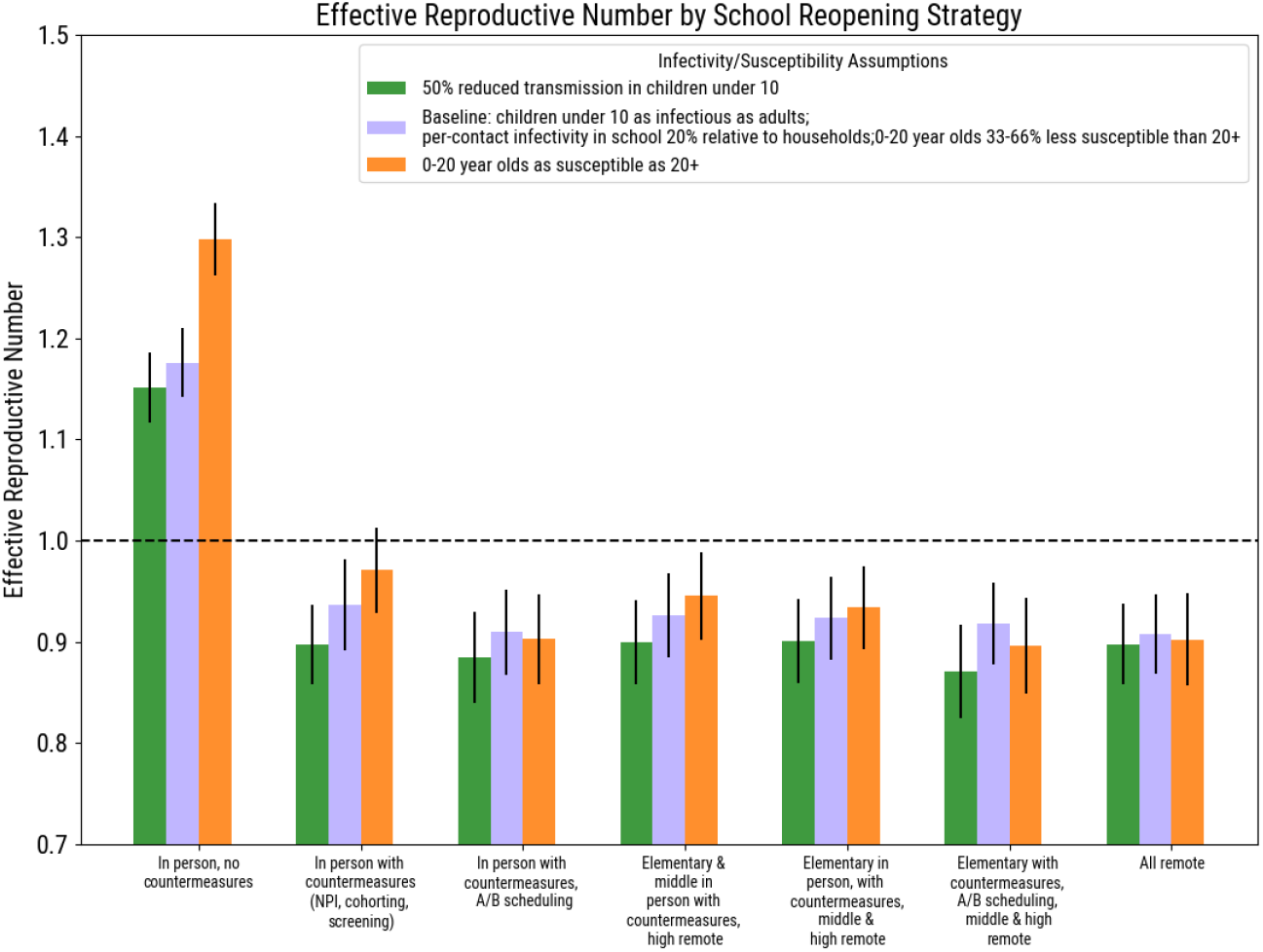
Effective reproduction number over the simulated period of school reopening (Sept 1^st^-Dec 1^st^) averaged across the top 20 parameter sets, assuming a COVID-19 case detection rate of 50 cases per 100,000 in the 14 days prior to school reopening. Error bars represent the standard deviation of the top 20 parameter sets.

### 3.1. Sensitivity analysis

Due to considerable uncertainty in the roles K-12 students, teachers, and staff play in COVID-19 transmission, we performed several sensitivity analyses to see if results were robust to a range of reasonable assumptions. While our baseline assumed that infected K-12 children are as infectious as adults, consistent with findings from PCR cycle threshold counts [12], a recent study using data from South Korea suggests that students under the age of 10 might be half as infectious as older students and adults [24]. Implementing this variation in the model leads to similar conclusions (Figure 6).

**Figure 6:**
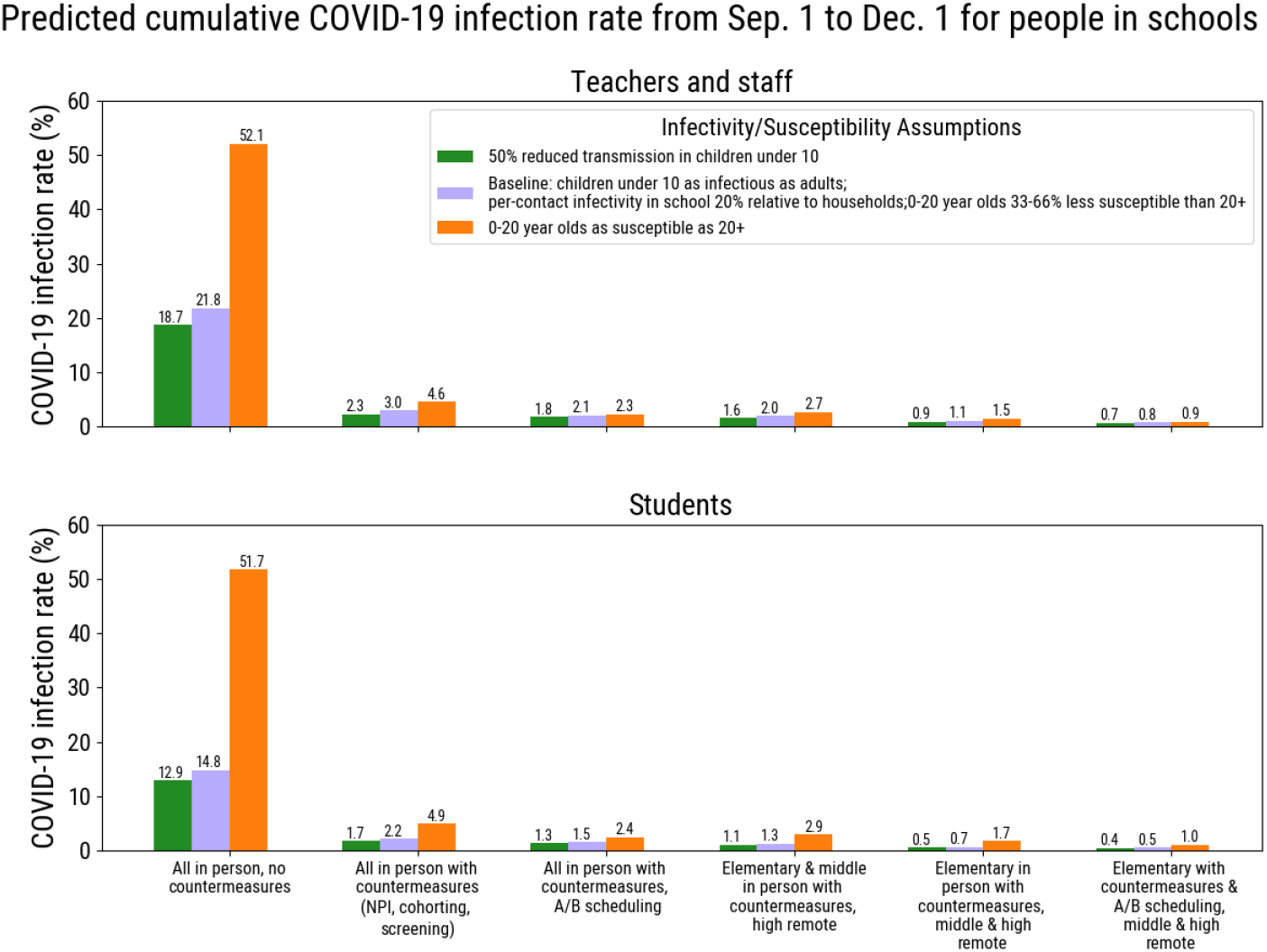
Cumulative COVID-19 infection rate for students, teachers and staff physically present in schools during the period of school reopening (Sept 1^st^-Dec 1^st^), averaged across the top 20 parameter sets, assuming a COVID-19 case detection rate of 50 cases per 100,000 in the 14 days prior to school reopening.

A second sensitivity analysis concerns the susceptibility of school-age children to COVID-19 infection. In our base case, we assume that 0-10 year-olds have a 66% reduced susceptibility to infection and 10-20 year-olds have a 33% reduced susceptibility to infection [31]. However, a recent study among children in Australia challenges this broadly held assumption, arguing that children are similarly vulnerable to the virus [14]. We find that without any countermeasures in schools, this assumption has a significant impact on the infection rate for people in schools (Figure 6) as well as the effective reproduction number in the community (Figure 5). With sufficient countermeasures, this assumption does not change our general policy conclusions.

## 4. Discussion

Any return to in-person learning will pose COVID-19 infection risk for students, staff and teachers. However, modeling results suggest that, depending on the case detection rate of COVID-19 in the community, a carefully organized incremental approach that returns the youngest students first with a reduced schedule would minimize the risk of infection within schools and provide important benefits to the neediest children. While this analysis is based upon school enrollment and population demographic data from King County, Washington, lessons can be applied to other settings to inform decision-making around school reopening. For example, as of August 11, 2020, King County had a 14-day case detection rate of 90 compared to 50 in New York City per 100,000 people. This would translate to nearly twice as many schools in the region with at least one person arriving at school with an active COVID-19 infection on September 1^st^compared to New York City, depending on the size of the school. It may also indicate a different optimal strategy for school reopening in these cities. In fact, Seattle Public Schools, citing a case detection rate in the city of well over 75 per 100,000 in the last two weeks of August, announced they would be starting the fall with completely remote learning [27], while New York Public Schools announced they would start with an A/B hybrid scheduling approach [30]. Schools in the San Francisco region have also mostly opted for remote learning, in line with recent model-based recommendations finding a high risk of school reopening [11].

We find that the solution with the lowest health risk also has the highest educational cost, in terms of days of distance learning required. We note that days of distance learning are not experienced equally by students and the benefit gained varies considerably based on both age and other factors such as socioeconomic status and resources available to their families [2]. Decision makers must therefore balance the benefits of in-person education with the safety of teachers and staff, while continuing efforts to decrease the incidence of COVID-19 outside of schools. Regardless of the approach taken, it will be critical to monitor schools with symptomatic screening and react quickly to the emergence of cases in schools with testing and contact tracing.

Our results support the strategy of returning elementary school students to school either full-time or with an A/B schedule while keeping all other students remote for the first three months of school to best minimize the risk of COVID-19 infection in schools. Returning elementary schools in-person first is both relatively lower-risk and higher-benefit; elementary school aged children are less susceptible to infection [31] and potentially less likely to transmit infection [24, 21]. Additionally, they may benefit more from in-person learning and pose more of a burden on family members. These results have been put into practice by some countries, which have returned elementary students to school first and older students at a later stage after observing an absence of significant school-based transmission [31, 1]. However, these countries had a significantly lower COVID-19 incidence rate prior to reopening schools than many parts of the US, including Washington State, are experiencing today.

### 4.1. Limitations

While agent-based modeling is able to capture many details of populations and disease transmission, our work has important limitations and assumptions that could impact our findings. There is still a high degree of uncertainty around the susceptibility, symptomiticity/severity, and infectivity of COVID-19 in children, particularly since schools in most locations shut down early in the epidemic. Our analysis is based on the most recent scientific literature for each of these parameters. We assumed individuals under 20 had a 45–50% reduced risk of developing symptoms [8] and 33–66% reduced risk of acquiring infection [31]. We varied this in sensitivity analysis so that individuals under 20 are equally as susceptible to infection as individuals age 20 to 50. We assume that an infectious individual is 5 times more likely per day to transmit to a household contact than a school contact, based on estimated numbers of hours spent in each setting per week. We assume all individuals infected with COVID-19 are equally likely to transmit infection per contact, and varied this assumption in sensitivity analysis so that individuals under 10 years old are half as likely.

After being diagnosed, all individuals are assumed to reduce their daily infectivity by 70% for home contacts, 90% for community contacts, and 100% for school and work contacts. Additionally, the household contacts of these individuals may be traced, notified, and school contacts removed from school for a full 14-day quarantine period. While we anticipate that schools will be able to help identify contacts of diagnosed students or staff, the large number of contacts within schools may place additional burden on local or state contact tracing efforts and our analysis does not represent this. We assume that household transmission is not reduced while children are attending school in person, nor that it increases when students are learning remotely.

In terms of structural choices, we made several assumptions about the implementation and impact of school reopening strategies. We chose to not model increased transmission associated with parents/guardians returning to work following a return to in-person learning. We also assume students who participate in remote learning are not in contact with anyone from school. On days that students are learning remotely, we do not account for any potential interaction students may have in a congregate care setting, such as an alternative after-school care program. We have assumed that, if implemented, all elementary and middle schools will be able to enforce classroom cohorting, whereby students are grouped into a classroom and are only in contact with other students and teachers in that classroom. We note that cohorting may be difficult to implement given the complexity of class scheduling for student bodies with multiple academic tracks, elective classes, and degree requirements. Therefore, we may be underestimating the transmission impact of school reopening, and further caution should be taken in any decisions to return to in-person learning.

We assumed that, if implemented, symptomatic screening would occur daily in schools and students or teachers presenting with COVID-like symptoms would be sent home. Those who are symptomatic may be asked to take a diagnostic test, which we assumed returns a result within two days. Students who received a negative test result return to school the next day, and students who received a positive test result are isolated at home for 14 days. Our results do not depend on school staff administering the diagnostic tests. We are not explicitly modeling after-school care, which many working parents depend upon to cover the gap between school hours and working hours. Families who use these services may also be more likely to be essential workers. We also do not model transportation to and from school, which may be an important source of transmission and which also depends on school resources.

## 5. Conclusions

Schools around the world are grappling with the challenge of returning to in-person learning in the COVID era. Much remains unknown about the role children play in COVID-19 transmission within schools and in the broader community, but the latest science suggests that younger children are less susceptible and show fewer symptoms if infected. From schools that never closed in Sweden to reopening examples in Europe and Asia, lessons on how the US might resume in-person learning are abundant and diverse. Our computational modeling synthesizes this evidence, and the latest results give reason for optimism.

Yet reopening schools is not a zero-risk activity. Symptom screening is imperfect, and COVID-19 will be present in the respiratory system of students, teachers, and staff on day one. Additionally, a return to in-person learning would allow parents and guardians return to work, which could be accompanied by an associated increase in transmission outside schools. But the solution with the lowest health risk has the highest educational cost, the majority of which lands on those families most marginalized and under-resourced: those without access to technology and private tutors, and whose parents or guardians work in the essential economy. Schools must open; the question is when and how, so as to balance the benefits of in-person education with the safety of teachers and staff, all while realizing that COVID is not just a school problem.

## Data Availability

The model used is available open source; all data are available from the authors upon request.

https://covasim.org

## Appendix A. Schools network structure

We simulated a representative sample of the 2.25 million King County, Washington residents in Covasim. We use student enrollment data for King County available from the 2018 American Community Survey [3]. This data gives an estimate of the enrollment rates by age for students attending any educational institution at the county level. Enrollment rates are given by age groups: ages 3–4, 5–9, 10–14, 15–17, 18–19, 20–24, 25–34, and 35–50. The enrollment rate for the last group is estimated to be less than 3%. School sizes are drawn from a distribution based on the enrollment numbers for Seattle area schools available for the year 2017 [26].

In order to model realistic school reopening scenarios, we equipped the model to generate networks within schools that reflect proposed cohorting by age and grade. We modeled schools to approximate age mixing patterns [10] between students within preschool, elementary schools, middle schools, high schools, and universities. Using the county-level school enrollment data [3], we simulate contacts within schools, mixing between students and teachers, and clustering of students into cohorts. Mixing of students and teachers can be thought of as following three main patterns: (1) students sorted in classroom cohorts of the same grade with one or two teachers, (2) students mixing with random contacts mostly within the same grade and at least one teacher, and (3) students mixing with random contacts across the entire school and at least one teacher. The first mixing pattern resembles the contact structures commonly found in pre-school and elementary schools, where students are generally taught by one teacher and stay with the same classroom of contacts throughout the day. The second pattern reflects mixing patterns often found in middle schools and high schools where students have individualized schedules and mostly interact with other students in the same grade. The third mixing pattern reflects university settings where student interaction occurs in classes, dorms, and in other spaces on campus. Student mixing in these institutions display less age assortativity because of the high variability of age when students enroll, use of common spaces such as libraries and dining halls, and other aspects of on-campus life.

In addition to students and teachers, schools also include additional staff members such as principals, counselors, nurses, maintenance, and cleaning staff. Using information on the estimated ratio of students to all staff members, we model the number of additional non teaching staff expected for each school and the contacts for them as random contacts across the entire school. This reflects the overall more varied contact patterns of non-teaching staff with students, teachers, and other staff members.

## Appendix B. Calibration

We used an optimization procedure to find a set of parameters that resulted in our desired combinations of effective reproduction number in the absence of school reopenings (specifically, *R*_e_ = 0.9) and cumulative diagnosed cases in the two weeks prior to the beginning of the school year (specifically, 20, 50, or 110 diagnosed cases per 100,000 people). The parameters used for the optimization were the number of seed infections at the start of the simulation, and the reduction of workplace and community contacts.

We calibrated these model parameters using the Tree-structured Parzen Estimator sampler in Optuna, a Python-based optimization library. The sampler finds regions of the parameter space that minimize a scalar objective function by learning the relationship between the parameters and the objective, as well as the overall probability of various objective values. We defined the objective function to be the sum of squared differences between the target values (i.e., average effective reproduction number from September 1st to December 1st; cumulative COVID-19 cases in the two weeks prior to September 1st) and the corresponding model outputs.

In all cases, we found a roughly 40% reduction in workplace and community contacts resulted in *R*_e_ = 0.9. Initializing the simulation with roughly 80, 160, and 400 seed infections resulted in scenarios of 20, 50, or 110 diagnosed cases per 100,000 people over a two-week period. We ran the analyses with the top 20 best-fitting parameter sets to each of these three scenarios.

